# Antibody and memory B-cell immunity in a heterogeneously SARS-CoV-2 infected and vaccinated population

**DOI:** 10.1101/2022.02.07.22270626

**Authors:** Eva Bednarski, Perla M Del Rio Estrada, Justin DaSilva, Celia Boukadida, Fengwen Zhang, Yara A Luna-Villalobos, Ximena Rodríguez-Rangel, Elvira Pitén-Isidro, Edgar Luna-García, Dafne Díaz Rivera, Dulce M López-Sánchez, Daniela Tapia-Trejo, Maribel Soto-Nava, Myriam Astorga-Castañeda, José O Martínez-Moreno, Guadalupe S Urbina-Granados, José A Jiménez-Jacinto, Francisco J Serna Alvarado, Yerania E Enriquez-López, Oliva López-Arellano, Gustavo Reyes-Teran, Paul D. Bieniasz, Santiago Avila-Rios, Theodora Hatziioannou

## Abstract

Global population immunity to SARS-CoV-2 is accumulating through heterogenous combinations of infection and vaccination. Vaccine distribution in low- and middle-income countries has been variable and reliant on diverse vaccine platforms. We studied B-cell immunity in Mexico, a middle-income country where five different vaccines have been deployed to populations with high SARS-CoV-2 incidence. Levels of antibodies that bound a stabilized prefusion spike trimer, neutralizing antibody titers and memory B-cell expansion correlated with each other across vaccine platforms. Nevertheless, the vaccines elicited variable levels of B-cell immunity, and the majority of recipients had undetectable neutralizing activity against the recently emergent omicron variant. SARS-CoV-2 infection, experienced prior to or after vaccination potentiated B-cell immune responses and enabled the generation of neutralizing activity against omicron and SARS-CoV for all vaccines in nearly all individuals. These findings suggest that broad population immunity to SARS-CoV-2 will eventually be achieved, but by heterogenous paths

## Introduction

The emergence of SARS-CoV-2 and variants thereof has highlighted how viral immune evasion, and variable global access to vaccines can profoundly impact the course of pandemics. Thusfar, in the COVID19 pandemic, numerous SARS-CoV-2 vaccines based on a single, near-ancestral SARS-CoV-2 variant have been developed and differentially deployed around the world^1-5^. The immunogenicity and ability of the distinct vaccine platforms to prevent infection, and impact the clinical sequalae of SARS-CoV-2 infection has been variable^1-5^. Moreover, over time the ability of all SARS-CoV-2 vaccines to prevent infection has generally deteriorated as immune responses have waned and variants with increased transmissibility and immune evasiveness have displaced prior variants^6-14^.

While multiple components of the immune response to SARS-CoV-2 antigens likely contribute towards the effectiveness of vaccines in preventing infection and disease, the most definitive correlate of protection against infection is the titer of neutralizing antibodies^15,16^. Moreover, the path that SARS-CoV-2 spike evolution has followed has clearly indicated that neutralizing antibodies have imposed selective pressure on viral populations^17-20^. A corollary of this observation is that that recently emerging SARS-CoV-2 variants, in particular B.1.1.529(omicron), have substantial degree of resistance to neutralizing antibodies elicited by earlier variants and B.1 based vaccines^6,10-15^. The majority of studies on SARS-CoV-2 vaccine elicited neutralizing antibody responses have focused on single vaccines corresponding to those distributed in high income countries. However, in low- and middle-income countries, vaccine deployment is far less uniform and it is therefore important to determine levels of immunity following administration of vaccines that represent those deployed globally^21,22^. Such data should help inform policy recommendations regarding additional vaccination doses and non-pharmaceutical interventions to mitigate disease burden.

Here, we measured antibody and memory B-cell immunity in a sample population comprising 197 vaccinated individuals in Mexico, a middle-income country in which five different vaccines were deployed in 2020-2021. We measured spike-binding and neutralizing antibody titers as well as the numbers of spike-specific memory B cells in recipients of each of the five vaccines. We also determined the ability of vaccine recipient plasma to neutralize emergent SARS-CoV-2 variants that have circulated in Mexico, including B.1.1.529(omicron), as well as an earlier variant B.1.1.519, originally detected in Mexico^23,24^. Our data reveal significant vaccine-dependent differences in the generation of spike binding and neutralizing antibodies and the generation of B-cell memory in uninfected and infected vaccine recipients, but suggest that broad immunity to SARS-CoV-2 variants can be achieved by heterogenous routes.

## Results

### SARS-CoV-2 genomic surveillance and vaccination in Mexico

Like many countries, Mexico has experienced successive waves of SARS-CoV-2 infection and the dominance of distinct variants over time. To determine the spectrum of SARS-CoV-2 variation that was present in Mexico during the study, we analyzed viral genomes reported between February 2020 and January 2022 to the Global Initiative on Sharing Avian Influenza Data (GISAID)^25^ (Fig. 1A). A total of 48,221 genome sequences were obtained from all 32 states of Mexico, with disparities in regional coverage (Fig. 1B). From February to May 2020, the ancestral B.1 and various derivatives thereof dominated but were largely displaced in the winter of 2020-2021 by B.1.1.519, a variant that was first described and achieved high prevalence in Mexico, but did not spread globally, unlike the contemporary B.1.1.7(alpha) variant (Fig. 1A). Indeed, importation of B.1.1.7(alpha) and P.1(gamma) were partly responsible for the displacement of B.1.1.519 in the late spring early summer of 2022, while a very few B.1.351(beta) sequences were also detected during that time. Thereafter, all these variants were displaced by B.1.617(delta) which was the dominant variant from July to November 2021. As has been the case in numerous other countries, the currently emergent, highly transmissible and antibody resistant B.1.1.529(omicron) variant displaced B.1.617(delta) during the winter of 2021-2022 (Fig. 1A).

**Figure 1.**
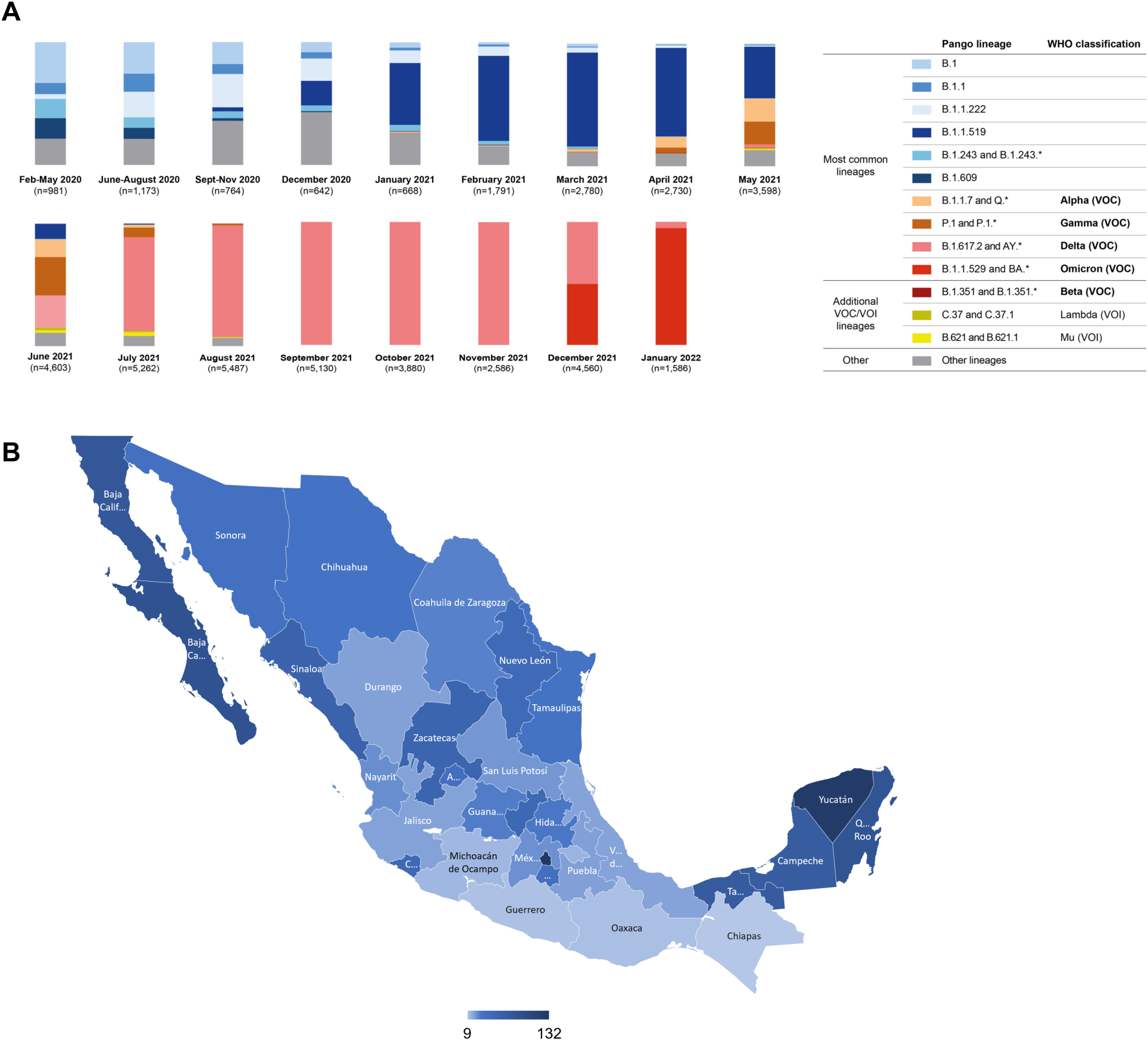
Frequency of SARS-CoV-2 variants in Mexico between February 2020 and January 2022. A total of 48,221 viral genome sequences obtained from samples collected in Mexico and downloaded from GISAID on January 28^th^, 2022, were analyzed. The frequency of variants was estimated over periods of a few months or individual months based on numbers of complete genomes sequenced. Most common lineages include variants circulating above 10% nationally in at least one period. In addition, less common lineages classified as variants of concern (VOC) and variants of interest (VOI) by the World Health Organization (WHO) were also included. Additional non-VOC/VOI lineages which circulated below 10% nationally during all periods were aggregated into the “Other” category. **(B)** Geographical distribution of viral genomes obtained in Mexico between February 2020 and January 2022. The number of genome sequences per 100,000 persons in the 32 states of Mexico is represented by a color gradient.

From December 2020 to spring 2021, five different vaccines were deployed in Mexico, specifically the BNT162b2 mRNA vaccine (Pfizer, 2-dose), as well as adenovirus-based vaccines ChAdOx1-S (AstraZeneca, 2-dose), Sputnik V (Gamaleya, 2-dose) and Ad5-nCoV (CanSino, single dose). Finally, an inactivated whole virion vaccine, CoronaVac (Sinovac, 2-dose) has been used in some locales. To date, these vaccines constitute 98% of the vaccine doeses administered in Mexico. We obtained blood samples from 197 individuals within 0.5 to 4.7 months post-vaccination. Of note, 80 (40.6%) of all participants had been infected with SARS-CoV-2 prior to collection of the blood sample. For 55 participants, infection was documented by a prior positive PCR test in 2020 or 2021, a median of 3.5 months (range 1 week-12 months) prior to vaccination (Sup Fig. 1). While the variant with which these individuals were infected was not determined, the B.1 and B.1.1.159 variants were most prevalent at the times at which PCR diagnoses were made (Fig. 1 and Sup Fig. 1). A further 25 participants had a positive test result for antibodies against the viral nucleocapsid (N) protein at the time of sampling (mRNA and adenovirus vaccine recipients only) but the time of infection relative to vaccination could not be determined. Participants without positive PCR diagnoses and negative anti-N antibody test were assumed to not have previously been infected. In the case of the CoronaVac recipients, the absence of prior infection could not be unequivocally established due to the presence of vaccine-elicited anti-N antibodies. For this group, participant self-report was used to assign prior absence of infection, but it is possible that some CoronaVac recipients had an undiagnosed infection prior to vaccination. The number of individuals that received each vaccine were: 29 BNT162b2, 38 ChAdOx1-S, 57 Sputnik V, 42 Ad5-nCoV and 31 CoronaVac.

### Comparison of plasma neutralization potency and breadth elicited by five SARS-CoV-2 vaccines

To compare the ability of each vaccine to elicit neutralizing antibodies, we employed neutralization assays^26^ using pseudoviruses carrying spike proteins derived from the ancestral SARS-CoV-2 variant (B.1) and variants that subsequently emerged in Mexico (Fig. 1). The range of plasma neutralizing titers elicited in previously naïve participants varied markedly among the different vaccines (Fig. 2). Specifically, in naive participants, the BNT162b2 mRNA vaccine elicited the highest overall 50% neutralizing titers (NT_50_) against the ancestral B.1 variant that closely matches the vaccine antigen (median = 2836). All of the previously uninfected participants that received the 2-dose BNT162b2 vaccine had detectable neutralization titers (range = 390-7651) (Fig. 2A). In contrast, previously uninfected individuals who received the CoronaVac vaccine had the lowest median neutralization titers against B.1 (median NT_50_ = 469, range = <50-3900), i.e. 6-fold lower than the BNT162b2 vaccine recipients. For the adenovirus-based vaccines, the single dose Ad5-nCoV vaccine gave NT_50_ values that were similar to CoronaVac (median NT_50_ = 542, range <50-1928) while the 2 dose adenovirus vaccines (ChAdOx1-S and Sputnik V) gave intermediate titers (median NT_50_ = 705, range 87-6169 and NT_50_ = 1013, range 190-4815) respectively. (Fig. 2A).

**Figure 2.**
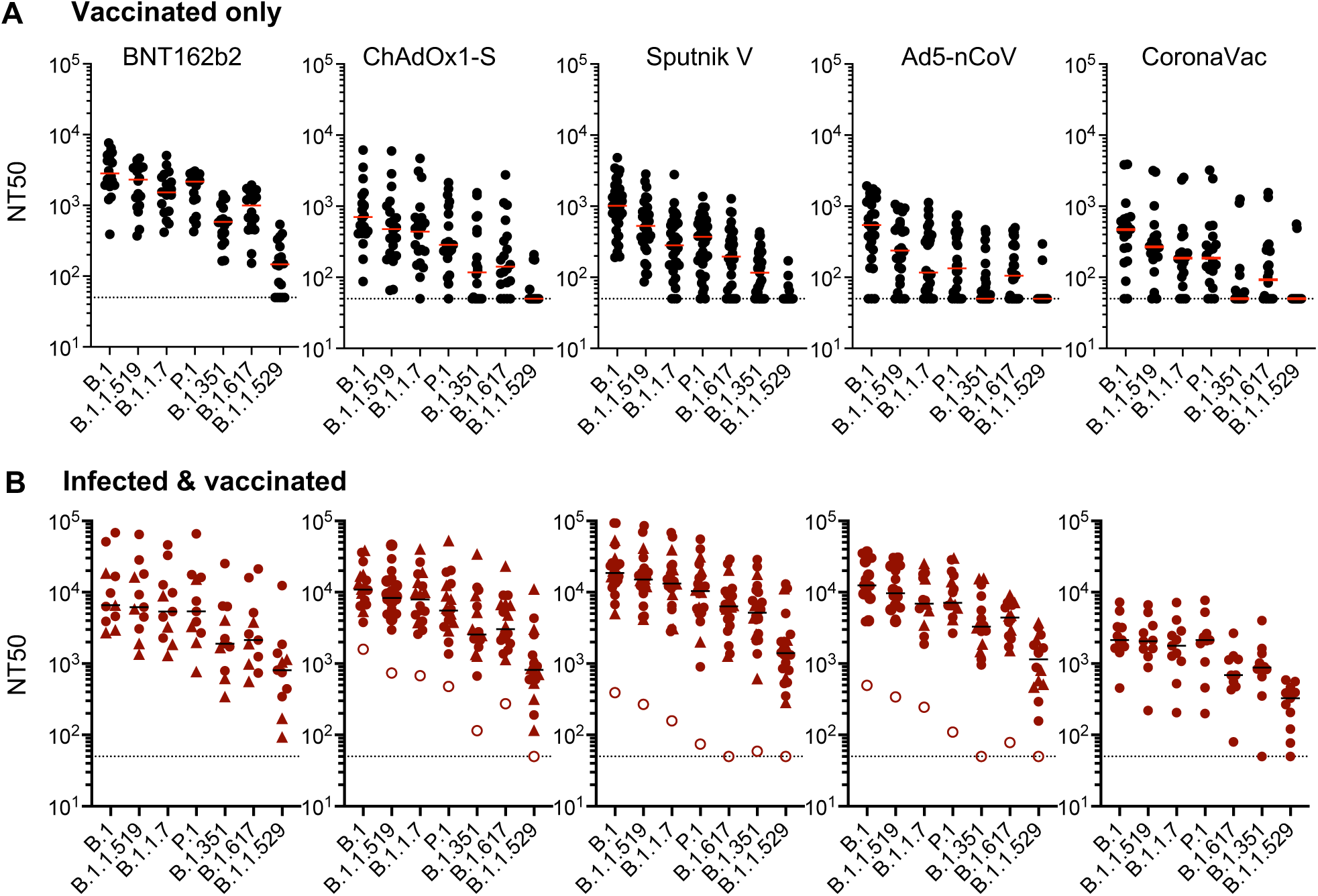
Plasma neutralization activity against SARS-CoV-2 variants in vaccine recipients. NT_50_ values of plasmas from recipients of one of five SARS-CoV-2 vaccines against B.1 or other SARS-CoV-2 variants. **(A)** Recipients with no documented prior infection with SARS-CoV-2. **(B)** Recipients were infected with SARS-CoV-2 prior to the study as documented by a PCR positive test (closed circles) or at an unknown time prior to sample collection as indicated by presence of anti-N antibodies (closed triangles). Individuals with prior positive PCR tests but seronegative for anti-N are indicated by open circles. The median values of 2-4 independent experiments for each plasma is plotted. Dashed line indicates the lowest plasma dilution tested (1:50). Lines indicate group median NT_50_ values.

For all the deployed vaccines, the neutralization potency of recipient plasma was progressively reduced for variants that subsequently dominated the SARS-CoV-2 viral populations in Mexico. For the B.1.1.519, B.1.1.7(alpha) and P.1(gamma) variants the reduction in potency was comparatively modest, NT_50_ values were 1.2- to 4.6-fold reduced compared to those against B.1 (Fig. 2A). Neutralization potencies against the subsequently emergent, more neutralization resistant variants were further eroded for all vaccine recipients with reductions in NT_50_ values ranging from 4.8- to 10.8-fold for B.1.351(beta) and 2.8- to 5.1-fold for B.1.617(delta). Indeed, for several vaccines, NT_50_ values against these variants fell below the limit of detection for a large fraction of participants. This was most noticeable for the CoronaVac vaccine, for which 13/20 and 9/20 plasma samples lacked detectable neutralizing activity against B.1.351(beta) and B.1.617(delta) respectively (Sup Fig. 2).

The recently emergent B.1.1.529(omicron) variant had the most substantial degree of neutralization resistance. Indeed, the majority (5/17, 16/20, 30/33, 24/26 and 18/20) of plasmas from uninfected BNT162b2, AstraZenca, Sputnik, Ad5-nCoV and CoronaVac recipients, respectively, had undetectable neutralizing activity against B.1.1.529(omicron) (Fig. 2A and Sup Fig. 2). Moreover, in the case of the uninfected CoronaVac recipients, these numbers may over-estimate the frequency of B.1.1.529(omicron) neutralizers (see below), as individuals with an undiagnosed prior infection could not be excluded from this group.

### Infection increases neutralizing antibodies for all SARS-CoV-2 vaccines tested

Prior work has shown that the titers and breadth of neutralizing antibodies elicited following vaccination were increased in individuals who were infected with SARS-CoV-2 months before receiving an mRNA vaccine^18,27-30^. We determined whether this potentiating property of infection in enhancing vaccine elicitation neutralizing antibody titers was generalizable. Of the 197 participants in our cohort, 80 had been infected as well as vaccinated, the majority of these (55 participants) were diagnosed by PCR in 2020 and early 2021, prior to vaccination, while the remaining 25 participants were demonstrated to have anti-N antibodies at the time of sampling. Plasma neutralizing titers against the ancestral B.1 variant in these infected and vaccinated individuals compared to the corresponding uninfected groups were increased for all vaccines. Specifically, the median (and range) NT_50_ values for infected BNT162b2, ChAdOx1-S, Sputnik V, Ad5-nCoV and CoronaVac recipients were 6593 (390-7651), 10893 (1591-38725), 18634 (391-93319), 12462 (494-36770) and 2133 (454-7259), representing values that were 2.3-fold to 23-fold higher than the uninfected counterpart groups (Fig. 2B). Infection also increased neutralizing antibody titers against the emergent variants as compared to the uninfected recipients for all vaccines (Fig. 2B). In contrast to uninfected vaccine recipients, nearly all infected and vaccinated individuals had detectable neutralizing activity against B.1.315(beta), B.1.617(delta) and B.1.1.529(omicron) variants. The median NT_50_ values against B.1.1.529(omicron) were 808 (93-12381), 816 (<50-11102), 1407 (<50-13025), 1144 (<50-3742) and 328 (74-589) for the infected BNT162b2, ChAdOx1-S, Sputnik V, Ad5-nCoV and CoronaVac recipients, respectively (Fig. 2B). Notably, 3 out of the 4 infected and vaccinated individuals that lacked detectable neutralizing activity against omicron were negative for anti-N antibodies at the time of sampling (open circles, Fig. 2B), suggesting low or absent prior SARS-CoV-2 antigen exposure, despite a prior positive PCR test.

Demographic characteristics (age and sex) had generally minor, non-statistically significant effects on neutralizing antibody titers. However, the number of individuals in each category was small (Sup Fig. 3A,B). Additionally, in the subset of individuals for which the time of prior infection documents by a PCR test was available, the time between infection and vaccination did not have a substantial effect on neutralization potency (Sup Fig. 3C).

### Broad neutralizing activity in infected and heterogeneously vaccinated individuals

We and others have previously reported that mRNA vaccination of individuals who were previously infected can generate neutralizing activity against distantly related sarbecoviruses such as SARS-CoV^6,18,27,28^. To assess the neutralization breadth of plasmas from infected individuals who received other vaccines, we measured neutralization titers against SARS-CoV (Fig. 3). Nearly all plasmas from infected vaccine recipients could cross-neutralize SARS-CoV with median (and range) NT_50_ values of 549 (105-6835), 1000 (258-5487), 1276 (238-5575), 1674 (362-3714) and 460 (183-1162) for the BNT162b2, ChAdOx1-S, Sputnik V and Ad5-nCoV and CoronaVac recipients, respectively (Fig. 3). Again, the 2 samples lacking SARS-CoV neutralizing activity were from the individuals that lacked anti-N antibodies while reporting a prior positive PCR test. As expected, titers against SARS-CoV were lower than those against SARS-CoV-2 B.1, but, interestingly, the neutralization potency of these plasmas against SARS-CoV, was comparable to that against B.1.1.529(omicron) despite the far greater divergence of SARS-CoV (Fig. 2B).

**Figure 3.**
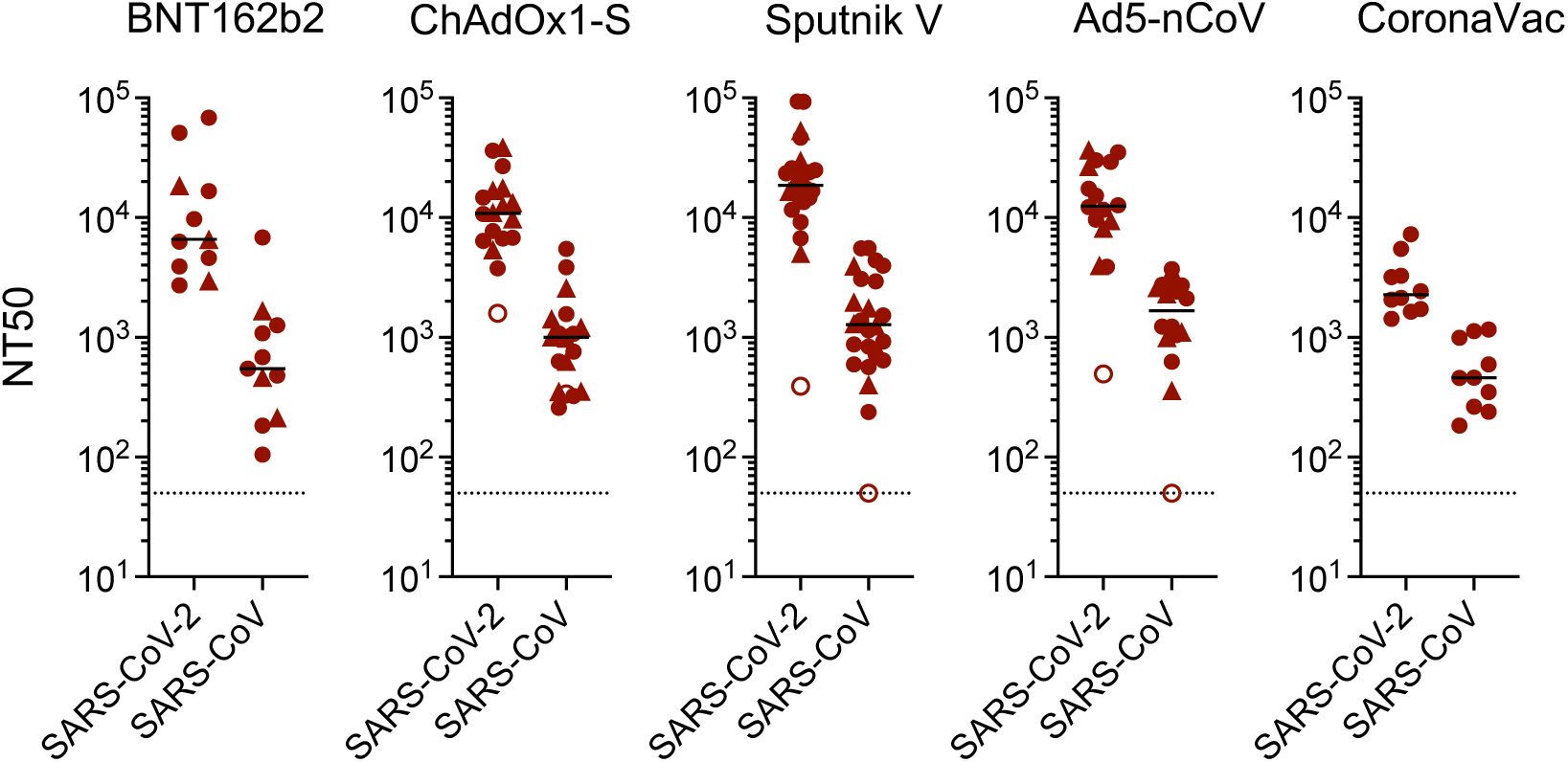
Plasma neutralization activity against SARS-CoV in in vaccine recipients. NT_50_ values measured in recipients who were infected with SARS-CoV-2 either prior to the study as documented by a PCR positive test (closed circles) or at an unknown time prior to sample collection as indicated by presence of anti-N antibodies (closed triangles). Individuals with prior positive PCR tests but seronegative for anti-N are indicated by (open circles). The median of 2 independent experiments for each plasma is plotted. Dashed line indicates the lowest plasma dilution tested (1:50). Lines indicated group median NT_50_ values.

### Expansion of spike-specific memory B-cells varies among vaccines and correlates with neutralizing titers

While neutralizing antibodies are the only component of the immune response that can, in principle, provide sterilizing protection, anamnestic responses following infection of vaccine recipients may contribute to prevention of disease. Thus, we measured the percentage of B.1 spike-binding and RBD-binding memory B-cells in recipients of each of the five vaccines. In previously uninfected individuals, the percentage of Spike-specific memory B-cells was highest for BNT162b2 recipients with a median of 0.022% of all memory B-cells specific for Spike, while the lowest median percentage, 0.003%, was found in individuals that received the CoronaVac vaccine (Fig. 4A). The ChAdOx1-S, Sputnik V and Ad5-nCoV vaccines generated an intermediate number of Spike-specific memory B-cells, (median = 0.006%, 0.007% and 0.004%) respectively. The number of RBD-binding memory B-cells in uninfected vaccinated individuals also followed this trend (Sup Fig. 4A).

**Figure 4.**
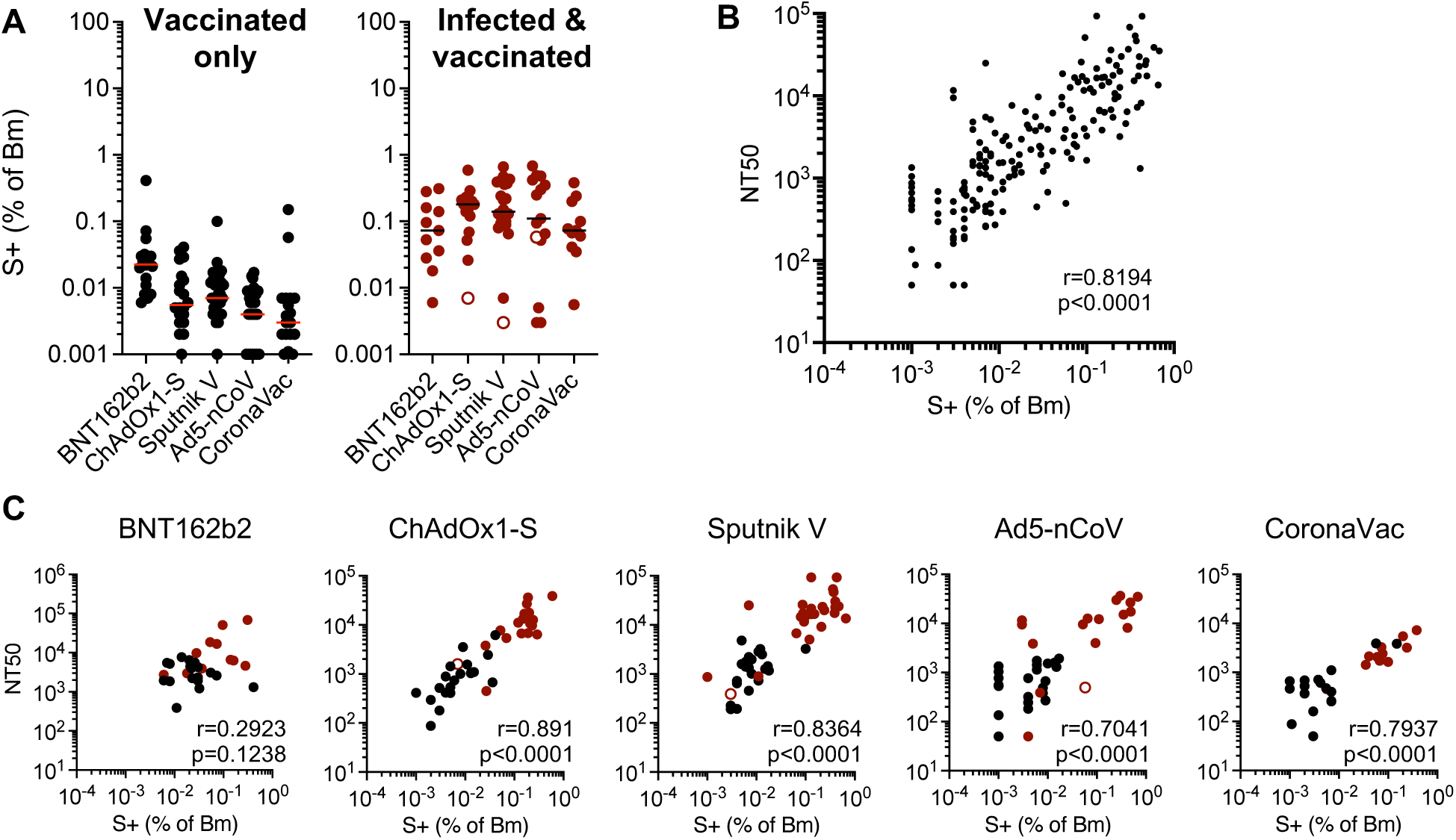
Quantification of SARS-CoV-2 spike specific memory B-cells in vaccine recipients. Memory B cells (Bm) in recipient PBMC were enumerated using FACS and a trimeric recombinant SARS-CoV-2 (B.1) spike protein. **(A)** Percentage of spike-binding (S+) Bm cells in recipients of one of five vaccines that were uninfected (left, black circles) or infected (right, red circles). (Open red circles = anti-N negative individuals as in Fig. 2B). Horizontal lines indicate the median percentage S+ (of Bm cells). **(B)** Correlation between neutralizing antibody titers (NT_50_) and percentage of S-binding Bm cells across all vaccine recipients **(C)** Correlation of neutralizing antibody titers (NT_50_) with percentage of S-binding Bm cells in each separate uninfected (black symbols) or infected (red symbols) vaccine recipient group. The r values indicate Spearman correlation coefficients

Infected and vaccinated individuals had a larger percentage of Spike-specific memory B-cells than those who were vaccinated but not infected with median values of 0.073%, 0.18%, 0.14%, 0.11% and 0.073% for BNT162b2, ChAdOx1-S, Sputnik V, Ad5-nCoV and CoronaVac recipients, respectively (Fig. 4A). Similarly, RBD-binding memory B-cells were greater in number for the infected versus uninfected vaccine recipients (Sup Fig. 4A). There was, overall, a general equalization of the numbers of memory B-cells in infected vaccine recipients compared to those who were vaccinated only, such that the differences between vaccine platforms were ameliorated (Fig. 4B). Notably, the degree to which Spike binding (and RBD-binding) memory B-cells were expanded in the blood of vaccine recipients correlated with the plasma neutralization titers in those same individuals (Fig. 4B, C and Sup. Fig. 4B). This finding was true for the overall dataset (Fig. 4B), and each individual vaccine (Fig. 4C) except for BNT162b2, where correlation did not reach statistical significance.

### Variable levels of antibodies that bind a conformationally intact spike are elicited by different vaccines and predict neutralizing activity and memory B-cell expansion

In addition to neutralizing antibodies, infection or vaccination induces antibodies that bind to Spike but do not neutralize. Such antibodies may contribute to the control of SARS-CoV-2 replication in vivo^31^. To measure spike binding antibodies, we developed a rapid and convenient assay that specifically measures antibodies that bind to a prefusion Spike trimer^19^. Specifically, we generated S-6P-NanoLuc, a secreted and conformationally stabilized (HexaPro) form of the SARS-CoV-2 B.1 Spike protein with NanoLuc luciferase fused at its C-terminus. We measured the amount of S-6P-NanoLuc luciferase activity that bound to participant derived immunoglobulins captured on Protein-G magnetic beads. In the uninfected subset of the cohort, the BNT162b2 vaccine elicited the highest Spike-binding antibody titers, with median S-6P-NanoLuc binding titers of 0.97×10^6^ relative light units (RLU)/μl of plasma, while the lowest levels of Spike binding antibodies were elicited by CoronaVac (median = 0.1×10^6^ RLU/μl) (Fig. 5A). The adenovirus vaccines elicited intermediate levels of Spike binding antibodies (median 0.16-0.2×10^6^ RLU/μl). As was the case for neutralizing antibodies and Spike-binding memory B-cells, infected and vaccinated participants generated higher levels of spike binding antibodies (median RLU/μl = 2.2, 2.1, 3.1, 2.5 and 0.54 ×10^6^ RLU/μl for BNT162b2, ChAdOx1-S, Sputnik, Ad5-nCoV, and CoronaVac recipients (Fig. 5A). The levels of Spike-binding antibodies correlated with the neutralizing antibody titers and Spike specific memory B cell expansion across vaccine platforms (Fig. 5B, C). Correlation between prefusion spike binding and neutralizing titers was evident for each vaccine tested and the quantitative relationship between spike binding antibodies and neutralizing antibodies was similar for each vaccine platform (Fig 5D). Notably, the levels of antibodies that bound the B.1 spike-based S-6P-NanoLuc also predicted neutralizing activity against B.1.617(delta), B.1.1.529(omicron) and SARS-CoV (Sup Fig. 5 A,B,C), even though the quantitative relationship between B.1 spike binding and neutralizing antibodies against other variants and SARS-CoV was different.

**Figure 5.**
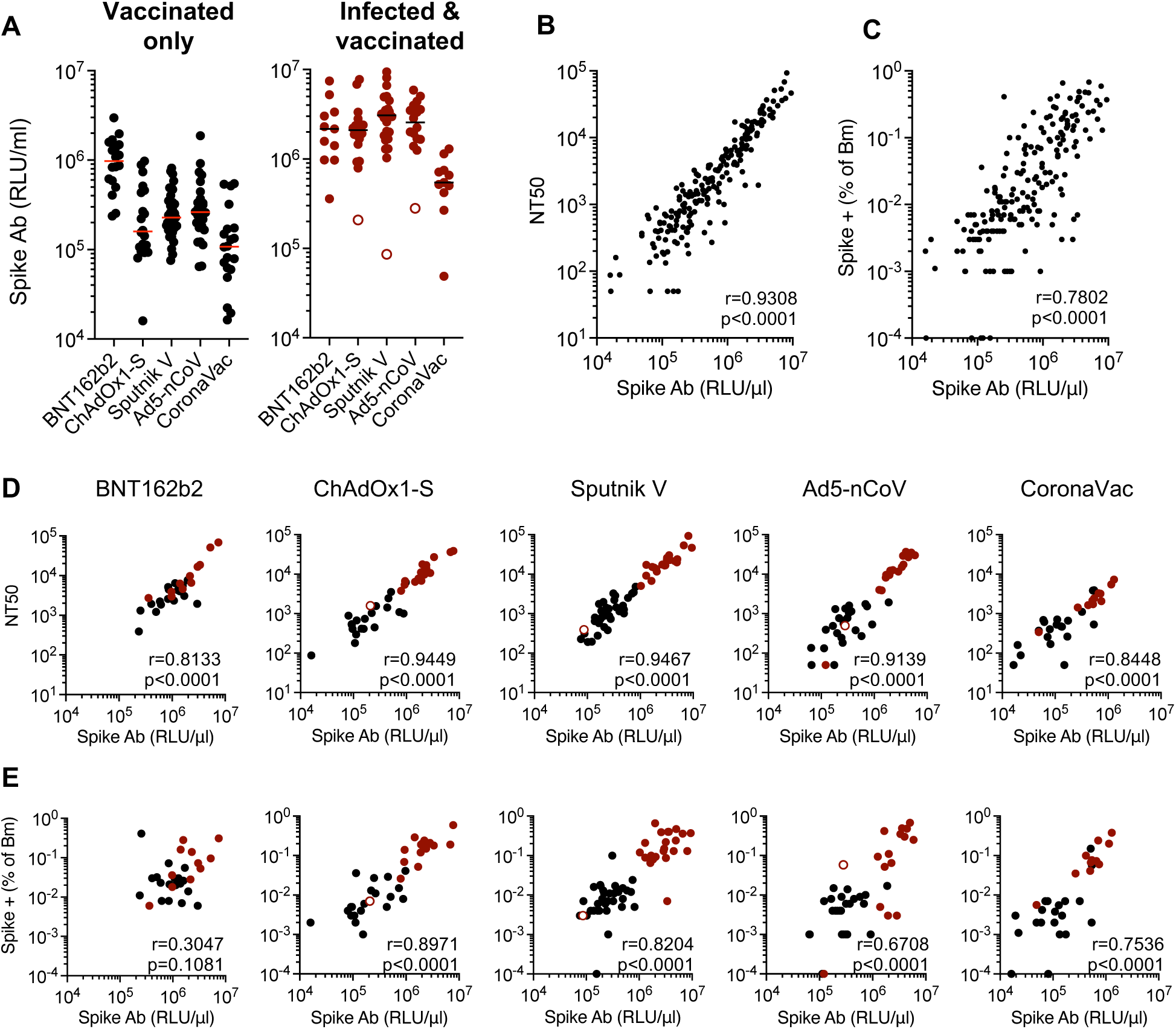
Quantification of antibodies that bind a prefusion SARS-CoV-2 spike protein. A conformationally stabilized trimer of a fusion protein between Spike (B.1) and NanoLuc (S-6P-NanoLuc) was used to measure Spike-binding antibodies. Antibodies in serially diluted participant plasmas were captured using protein G magnetic beads, then incubated with S-6P-NanoLuc and bound NanoLuc activity quantified. **(A)** Captured NanoLuc activity expressed as Relative Light Units (RLU) per μl of plasma from uninfected vaccine recipients (left, black circles) or infected (right, red circles). (Open red circles indicate anti-N-negative samples as described in Fig. 2B). Mean of two independent experiments. Lines indicate group median RLU/μl. **(B, C)** Correlation between neutralizing antibody titers (NT_50_) (B) or Spike specific memory B-cell expansion (C) and Spike-binding antibodies (RLU/μl) across all vaccine recipients. **(D, E)** Correlation between neutralizing antibody titers (NT_50_) (D) or spike specific memory B-cell expansion (E) and Spike-binding antibodies (RLU/μl) for each uninfected (black) or infected (red) vaccine recipient group. The r values indicate Spearman correlation coefficients

Spike-binding antibody levels also correlated with the numbers of Spike- and RBD-specific memory B-cells for all vaccines, though the correlation for BNT162b2 did not reach statistical significance (Fig. 5E, Supplementary Fig. 5D). Overall therefore, different measurements of B-cell immunity, specifically Spike binding antibodies, neutralizing titers, and Spike specific memory B-cell expansion, correlated with each other across all vaccine platforms tested.

### Longitudinal variation in neutralizing activity

A caveat associated with the aforementioned comparisons between vaccine platforms was non-uniformity in the time between receipt of the vaccine and sample collection. We therefore determined the extent to which titers induced in vaccine recipients from all five vaccines used in this study changed over time against B.1, and the two most recently prevalent variants B.1.617(delta) and B.1.1.529(omicron). Most participants (170/197) returned for a follow up visit, 2.5-7.8 months after the first (Fig. 6).

**Figure 6.**
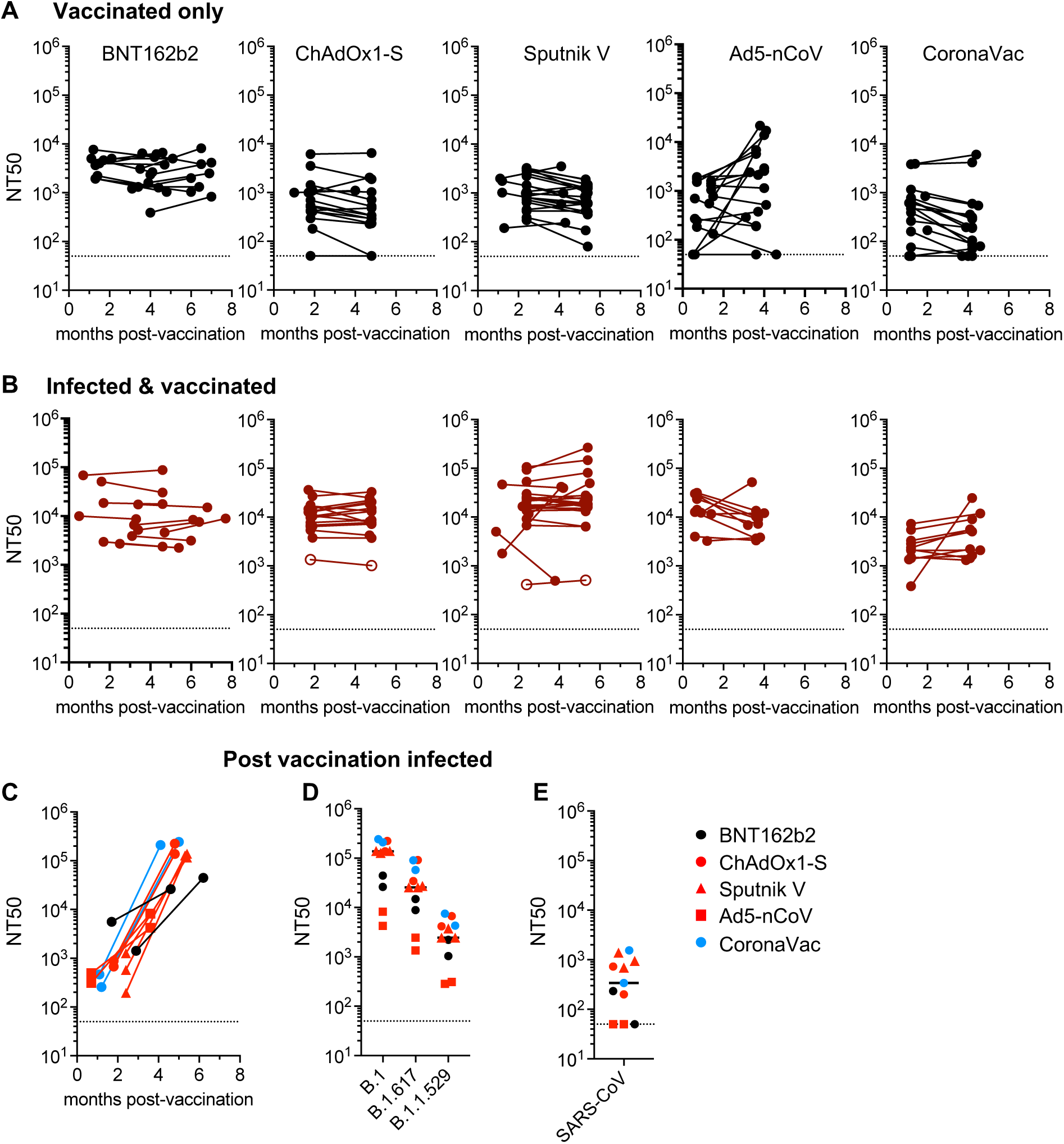
Longitudinal analysis of neutralizing antibodies post vaccination. Comparison of neutralizing antibody titers between the first and second plasma sample for each participant. **(A)** Uninfected participants **(B)** SARS-CoV-2 infected participants. (Open circles individuals that were seronegative for anti-N as in Fig 2B.) **(C)** Change in NT50 values for participants that were infected between the acquisition of the 2 samples as indicated by acquisition of, or large increase in, anti-N antibodies. **(D, E)** Neutralizing antibody titers against SARS-CoV-2 variants (D) and SARS-CoV (E) in second samples from participants who were infected between collection of the two samples. One sample from the Ad5-nCoV group with a prior positive PCR test and anti-N antibodies in the second but not the first sample was included in this group. The median of 2 independent experiments for each plasma is plotted. Dashed line indicates the lowest plasma dilution tested (1:50). Horizontal lines indicate group median NT_50_ values.

For most uninfected participants who remained anti-N negative at both visits, there were small differences in the NT_50_ values measured for B.1 at the second visit compared to the first (Fig. 6A). With the caveat that the timing intervals were not uniform among the vaccine platforms, BNT162b2 vaccine recipients displayed a marginal decrease (∼0.9-fold) in neutralizing titer at the second visit, while the recipients of ChAdOx1-S, Sputnik V and CoronaVac increased slightly ∼1.4-2.4-fold (Fig. 6A). Ad5-nCoV recipients showed a 4-fold increase in titers against B.1 at the second visit, though this might be due to the first visit samples being collected very early for the majority of this group, (Fig. 6A). The deficit in B.1.617(delta) and B.1.1.529(omicron) neutralization at the second visit was similar to that at the first visit for all vaccines (Sup Fig. 6A). As observed for the first visit, NT_50_ values in infected and vaccinated participants were higher than the uninfected groups (Fig. 6B and Sup Fig. 6B) and difference in median NT_50_ between infected and uninfected groups at the second visit was similar to that at the first. However, in the Ad5-nCoV recipients, the 25-fold difference in NT_50_ values for infected and uninfected groups at first visit was only 7-fold at the second visit (Fig 2A,B and Sup Fig. 6A,B). This discrepancy may be due to the comparatively short interval between vaccination and the first visit for this group, as discussed above.

### Effect of infection after vaccination on neutralizing antibody titer and breadth

Of the 78 returning mRNA or adenovirus vaccine recipients that were anti-N negative and assumed to be not infected prior to the first sample collection, 8 participants had anti-N antibodies in the second sample. A further 2/19 returning recipients of the CoronaVac whole inactivated vaccine exhibited an atypical increase in anti-N titers in the second sample. A single Ad5-nCoV recipient that was reported PCR positive but was anti-N negative at the first visit became anti-N positive at the second visit. These 11 participants were judged to be infected during the course of the study, after vaccination, and were considered separately. Based on the time of sample collections the majority of these post-vaccination infections occurred between June and September 2021. The B.1.617(delta) was the most prevalent variant during the majority of this period but we cannot exclude the possibility that B.1.1.519, B.1.1.7(alpha) and P.1(gamma) variants were responsible for post vaccination infections (Fig. 1). In all of these individuals, neutralizing titers increased considerably compared to the first visit (Fig. 6C) and had a median NT_50_ values of 138105 (4227-243267) or 43- to 710-fold higher than returning vaccine recipients who did not seroconvert with anti-N antibodies (mRNA and adenovirus vaccine recipients) or did not have increased anti-N titers at the second visit (CoronaVac recipients) (Fig. 6C, D). Plasmas from all 11 of these post-vaccination infected participants were able to neutralize B.1.617(delta) and B.1.1.529(omicron) with median (and range) NT_50_ values of 25523 (1349-91522) and 2457 (284-7511) respectively (Fig. 6D), values comparable to those exhibited by plasmas from infected-then-vaccinated individuals (Fig. 2B and Sup Fig 6B). Most of the vaccinated-then-infected plasmas were also able to neutralize SARS-CoV, but with potencies that were a median of 7-fold lower than that against B.1.1.529(omicron) (Fig. 6D,E).

## Discussion

The immune responses elicited by the SARS-CoV-2 mRNA vaccines were extraordinarily effective at providing short-term protection against infection by closely matched SARS-CoV-2 variants^32,33^. Subsequently, waning immunity and emergence of viral variants with greater transmissibility and antibody evasiveness, including in Mexico^23,24^, has eroded the ability of vaccines to prevent infection^6-14^. Most studies have focused on vaccines deployed in wealthier nations, and direct comparisons among vaccines available to less resource-rich countries are limited^21,22^. The data presented herein reveal important differences in the ability of vaccines employed in such countries to elicit neutralizing antibodies and B cell memory, in both naive and previously infected individuals and against multiple variants. The ChAdOx1-S, Sputnik V, Ad5-nCoV and CoronaVac vaccines investigated had variable abilities to elicit neutralizing antibodies, all generating lower titers than the BNT162b2 mRNA vaccines against all variants. They are therefore expected to exhibit lower levels of protection against infection.nIndeed, many previously uninfected vaccine recipients evaluated herein had undetectable levels of neutralizing antibodies against recently emergent SARS-CoV-2 variants. In particular, the majority of previously uninfected vaccine recipients lacked detectable neutralizing activity against omicron and were thus unlikely to be protected against infection.

The deployment of incompletely effective vaccines during an ongoing pandemic has the consequence that population immunity is currently accumulating through heterogenous routes. In one frequent scenario, infection by SARS-CoV-2 might be followed by administration of one of a number of vaccines. In another, vaccination might be followed by infection. In a third, an initial vaccination might be followed by boosting with a vaccine of a different type. Finally, in populations with poor vaccination distribution or uptake, immunity might be accrued through multiple infections. In the context of evolving SARS-CoV-2 variants the nature of exposures to SARS-CoV-2 antigens and levels of immunity is therefore highly variable between individuals.As we and others have previously reported for mRNA vaccines^18,27-29,34^, prior infection strongly potentiated the ability of each of the vaccines investigated herein to elicit neutralizing antibodies. Additionally, prior infection increased the ability of the vaccines to elicit and expand Spike and RBD-specific memory B-cells. Prior experience of SARS-CoV-2 antigens, and consequent affinity maturation of B-cells^17,27,35-37^ likely explains why nearly all infected-then-vaccinated participants had neutralizing antibodies against omicron and a divergent sarbecovirus SARS-CoV. A small number of the participants in this study immunized with each of the vaccines acquired infection post vaccination. All of these individuals also exhibited high neutralizing titers, approximately equivalent to those in infected-then-vaccinated individuals. Most individuals who were infected post vaccination also had the ability to neutralize omicron and SARS-CoV. Thus, combinations of vaccination and infection might have similar outcomes, irrespective of the order in which they occur. If so, vaccination and infection might progressively build and broaden population immunity and decrease overall disease burden.

While omicron has spread rapidly among populations with high levels of vaccination, it has nevertheless imposed a reduced disease burden than prior variants. Anamnestic B-cell and T-cell immune responses as well as non-neutralizing and neutralizing anti-spike antibodies likely contribute to reduced disease severity. Notably, our data demonstrate that a simple quantitative measurement of antibodies that bind to a stabilized prefusion conformation spike protein can predict levels of neutralizing antibodies, as well as the degree of spike-specific and RBD-specific memory B-cell expansion. Appropriate serological assays might therefore be useful in prognostication of the likelihood of mild versus severe outcomes as well as the likelihood of infection in a population. This in turn could inform health care strategies, for example in a resource poor setting, such as Mexico where multiple different vaccines were utilized, perhaps targeting deployment of booster shots to those that received vaccines that elicited low levels of antibodies. This information could also guide the use of vaccines that are not widely used in wealthier nations and broaden options when confronting inequity in vaccine distribution, production limitations as well as hesitancy issues.

## Data Availability

All data produced in the present work are contained in the manuscript

## Acknowledgements

We gratefully acknowledge GISAID data contributors, including authors from the originating laboratories responsible for obtaining the specimens and the submitting laboratories from which viral genome sequences were generated and shared via the GISAID Initiative. This work was supported by the Mexican Government (Programa Presupuestal P016; Anexo 13 del Decreto del Presupuesto de Egresos de la Federación) by NIH grants R37AI64003 and R01AI501111 (PDB).; R01AI78788 (TH) and P01AI165075 (PDB and TH)

## Methods

### Participants

Over the period of winter 2020 to spring 2021, the following vaccines became available and were administered to the Mexican population; one based on mRNA, BNT162b2 (Pfizer/BioNtech), three based on adenovirus vector platforms, ChAdOx1-S (AstraZeneca), Sputnik V (Gamaleya) and Ad5-nCoV (CanSino), and one based on whole inactivated virus, CoronaVac (Sinovac). Participants were recruited by the Centre for Research in Infectious Diseases of the National Institute for Respiratory Diseases (INER-CIENI) in Mexico City, in collaboration with municipal authorities of five of the Mayor’s Offices of Mexico City (Coyoacán, Cuajimalpa, Iztapalapa, Magdalena Contreras and Tlalpan). Each municipality contributed with participants vaccinated with one of the approved vaccines mentioned above, following the recommended doses and intervals, according to the national vaccination plan. All donors provided written informed consent in compliance with protocols set forth and approved by the Comité de Ética en Investigación and the Comité de Investigación (Research Ethics Committee and the Research Committee) from INER Institutional Review Board (study no. B01-21). Blood collection was performed by CIENI-INER staff at health centers of the participating Mayor’s Offices. Epidemiological data of participants was collected on the day of blood donation. Evidence of a positive SARS-CoV-2 test was requested from those participants that declared prior COVID-19 infection before vaccine application. Additionally, SARS-CoV-2 infection was evaluated in all participants by detection of anti-N antibodies in plasma by ELISA (Elecsys Anti-SARS-CoV-2. Roche, Cat No 09203095190).

Samples were collected between 0.5-4.2 months post-vaccination from the 197 individuals that had received one of the five different vaccines. Whole blood was collected in acid citrate dextrose (ACD) tubes and processed for peripheral blood mononuclear cells and plasma isolation. Plasma samples were obtained after centrifugation and stored at −80°C. Peripheral blood mononuclear cells (PBMC) were obtained from whole blood by Ficoll-Hypaque density gradient centrifugation and cryopreserved at −140°C.

Plasma samples were also collected at a follow-up visit for 170 participants that returned 2.5-7.8 months after the first visit. Anti-N titers were measured for all returning participants as above.

### Pseudotyped virus neutralization assays

SARS-CoV-2 pseudotyped particles were generated as previously described ^26^. Briefly, 293Tcells were transfected with pNL4-3ΔEnv-nanoluc and pSARS-CoV-2-SΔ19. At 48 hours later particles were harvested, filtered and stored at −80°C. The amino acid deletions and/or substitutions corresponding to SARS-CoV-2 varisnts were incorporated into a spike expression plasmid using synthetic gene fragments (IDT) or overlap extension PCR mediated mutagenesis and Gibson assembly. Specifically, the variant-specific deletions and substitutions introduced inot the B1 sequence were:

B.1.1.519: T478K/D614G/P681H/T723A

B.1.1.7 (alpha):ΔH69/V70/ΔY144/N501Y/A570D/D614G/P681H/T761I/S982A/D1118H

P.1 (gamma): L18F/T20N/P26S/D138Y/R190S/K417T/E484K/N501Y/D614G/H655Y

B.1.351 (beta): D80A/D215G/L242H/R246I/K417N/E484K/N501Y/D614G/A701V

B.1.617 (delta): T19R/ Δ156-8/L452R/T478K/D614G/P681R/D950N

B.1.1529 (omicron): A76V/ Δ69-70/T95I/G214D/ Δ143-145/ Δ211/L212I/ins214EPE/G339D/S371L/S373P/S375F/K417N/N440K/G446S/S477N/T478K/E484A/Q493K/G498R/N501Y/Y505H/T547K/D614G/H655Y/N679K/P681H/N746K/D796Y/N856 K/Q954H/N969K/L981F

All spike proteins used in the pseudotype neutralization assays had a 19 amino acid C-terminal deletion and included the R683G substitution, which disrupts the furin cleavage site and increases particle infectivity without grossly affecting antibody sensitivity.

Fivefold serially diluted plasmas from vaccinated individuals were incubated with SARS-CoV-2 pseudotyped virus for 1 h at 37 °C. The mixture was subsequently added to an HT1080-based cell line engineered to express human ACE2 (HT1080.ACE2 cl14). The starting serum dilution on cells was 1:50. Nanoluc Luciferase activity in lysates was measured 48 hours post-inoculation using the Nano-Glo Luciferase Assay System (Promega) with the Glomax Navigator (Promega). Relative luminescence units were normalized to those derived from cells infected with SARS-CoV-2 pseudotyped virus in the absence of serum. The half-maximal neutralization titers for sera (NT_50_) were determined using four-parameter nonlinear regression (least squares regression method without weighting; constraints: top=1, bottom=0) (GraphPad Prism) and median values calculated for each sample.

### Spike-binding antibody assay

A recombinant conformationally stabilized (HexaPro) and secreted (transmembrane and cytoplasmic domain deleted) form of the SARS-CoV-2 B.1 Spike protein was generated with NanoLuc luciferase, a 3CL protease site and 6xHis tag fused at its C-terminus (S-6P-NanoLuc). The S-6P-NanoLuc protein was expressed in 293-Expi cells, captured on Ni^2+^-magnetic beads and the purified protein eluted following incubation with 3CL protease. Plasma samples from vaccinated individuals were initially diluted 1:10 in 2% BSA/PBS and then serially diluted fivefold in the wells of a 96-well plate. Immunoglobulins were captured on protein G magnetic beads (Lytic Solutions) diluted in 2% BSA/PBS for 1 hour at 4°C. A purified human monoclonal antibody against SARS-CoV-2 Spike, C144, diluted 1:20 in 2% BSA/PBS and serially diluted, was used as a positive control. The immunoglobulin coated beads were washed twice with 2% BSA/PBS and mixed with 10ng of S-6P-NanoLuc. After a 2-hour incubation at 4°C, beads were washed twice and transferred to a fresh 96-well plate. 50μl of reporter lysis buffer (Promega) was added to the beads and antibody-captured NanoLuc luciferase activity measured using the Nano-Glo Luciferase Assay System (Promega) with the Glomax Navigator (Promega). Non-parametric Spearmen’s correlation analysis (GraphPad Prism) was performed and p values calculated with a two-tailed analysis with 95% confidence interval.

### Measurement of Spike binding memory B-cells

Quantification of B cells specific to SARS-CoV-2 S (& RBD) was performed using S (and RBD) proteins multimerized with fluorescently labeled streptavidins based on methods described in^30^. To multimerize SARS-CoV-2 proteins with fluorescently labeled streptavidins, biotinylated SARS-CoV-2 spike protein (Biolegend, Cat No 793806) was mixed with streptavidin PE (BD, Cat No 554061) and streptavidin Alexa Fluor 647 (Biolegend, Cat No 405237) at ∼6:1 molar ratio, and biotinylated RBD protein (Biolegend, Cat No 793906) mixed with streptavidin APC Cy7 (BD, Cat No 554063) at a ∼4:1 molar ratio. Biotinylated proteins were incubated with the corresponding streptavidin at 4°C for one hour. Streptavidin BV510 (Biolegend, Cat No 405234) was used as a decoy probe, free biotin from Avidin-Biotin Blocking System (Biolegend, Cat No 927301) was used to help avoid cross-reactivity of antigen probes. Cryopreserved cells were thawed and rested overnight in complete medium (RPMI-1640, 10% heat-inactivated fetal bovine serum, 1% penicillin, 1μg/ml streptomycin, and 1% L-glutamine) prior to staining. Ten million PBMC were first incubated for one hour at 4°C with a cocktail containing SARS-CoV-1 multimerized proteins (200ng of Spike protein, 100ng per probe; and 27.5ng of RBD protein). Free biotin was added to this cocktail (5 mL). Cells were then washed twice by adding 3 ml of Stain Buffer (BD, Cat No 554657) and centrifuged (1500rpm for10 minutes). Cells were stained with surface antibodies and viability marker (Aqua dye Fluorescent Reactive Dye, Invitrogen, Cat No L344957) as follows: cells were incubated with IgG BV786 (20 min incubation at 4°C, BD, Cat No 564230, clone G18-145) followed by a ten minute incubation (4°C) with viability marker and Fc block (Biolegend, Cat No 422302), and by incubation with a cocktail of surface antibodies containing: IgM BV605 (clone G20-127, BD Cat No 562977); CD19 BV421 (clone HIB19, BD Cat No 562440); CD27 PERCP CY5.5 (clone M-T271, BD Cat No 560612); IgD PECY7 (clone IA6-2, BD Cat No 561314); CD3 PECF594 (clone UCHT1, BD Cat No 562280). Cells were washed and fixed with 1% paraformaldehyde diluted in PBS. Stained PBMC were acquired in a BD FACS Fusion using FACS DIVA software (v8.0) and data were analyzed using FloJo v10.8.0 software. Frequency of SARS-CoV-2 S-specific memory B cells was expressed as a percentage of CD27 negative B cells. (Analysis strategy: single cells > lymphocytes (FSC vs. SSC) > live cells > CD3 negative > CD19 positive cells > CD27 negative cells. Frequency of SARS-CoV-2 RBD-specific memory B cells were measured as a percentage of S+ cells (gated inside the S positive cells). Non-parametric Spearman’s correlation analysis (GraphPad Prism) was performed and p values calculated using a two-tailed test.

**Supplementary Figure 1.**
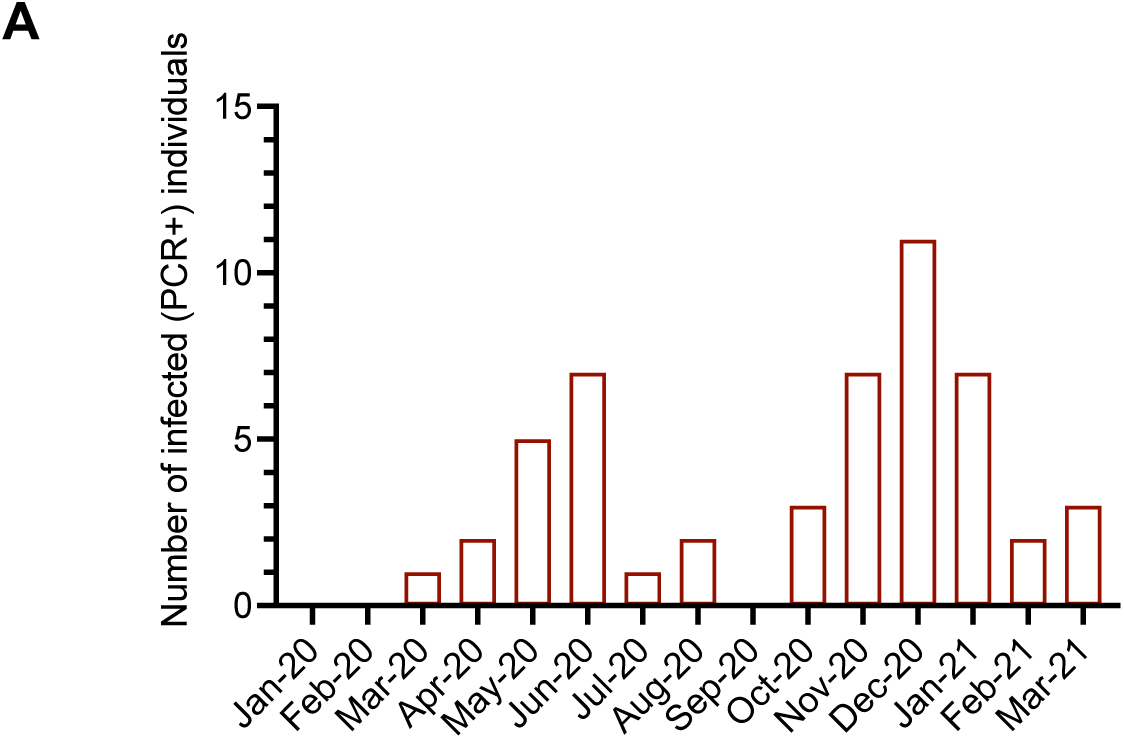
Distribution and participant infection over time. (**A)** Number of participants that were diagnosed as infected by PCR each month during January 2020 to March 2021.

**Supplementary Figure 2.**
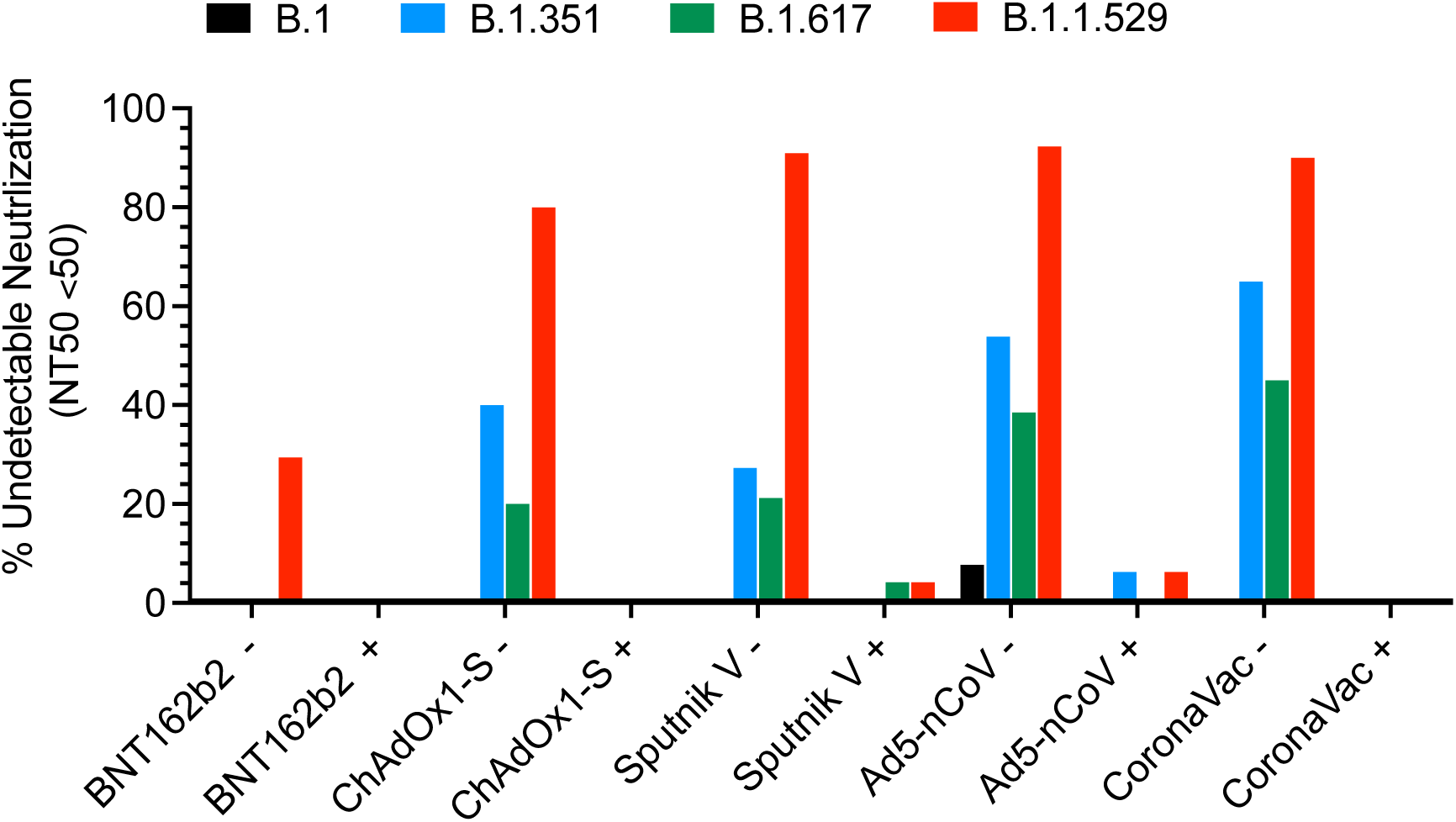
Fraction of participants with undetectable neutralizing antibody titers. The percentage of samples for each vaccine group that had NT50 below 50 (the lowest plasma dilution used) against B.1, B1.351(beta), B.1.617(delta) and B.1.1.529(omicron) was calculated for uninfected (-) and infected (+) participants.

**Supplementary Figure 3.**
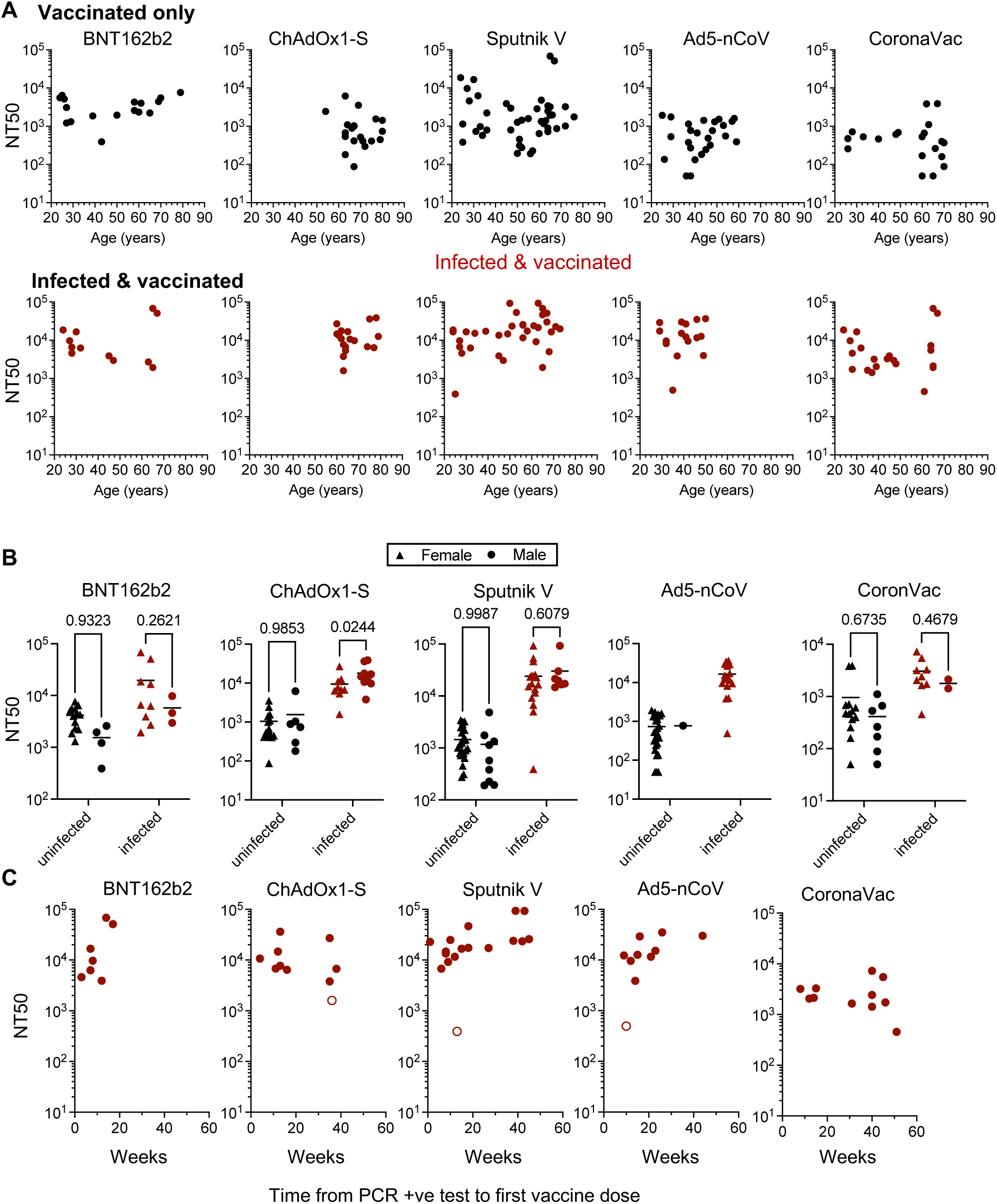
Correlation of participant characteristics and neutralizing antibody titers. **(A)** Participant age in each vaccine group and neutralizing antibody titers (NT_50_). Top (black circles), recipients with no documented prior infection with SARS-CoV-2. Bottom (red circles) recipients that were infected with SARS-CoV-2 either prior to the study or at an unknown time prior or between vaccination and sample collection. Participants with prior positive PCR tests but undetectable anti-N antibodies (open circles). **(B)** Participant sex in each uninfected (black) and infected (red) vaccine group and neutralizing antibody titers (NT_50_). p-values were calculated using a 2-way ANOVA Šidák’s multiple comparison test. **(C)** Time between infection (positive PCR test) and vaccination and neutralizing antibody titers (NT_50_). Only samples with available PCR tests were included in this analysis. Open circles indicate participants with prior positive PCR positive tests that were negative for anti-N.

**Supplementary Figure 4.**
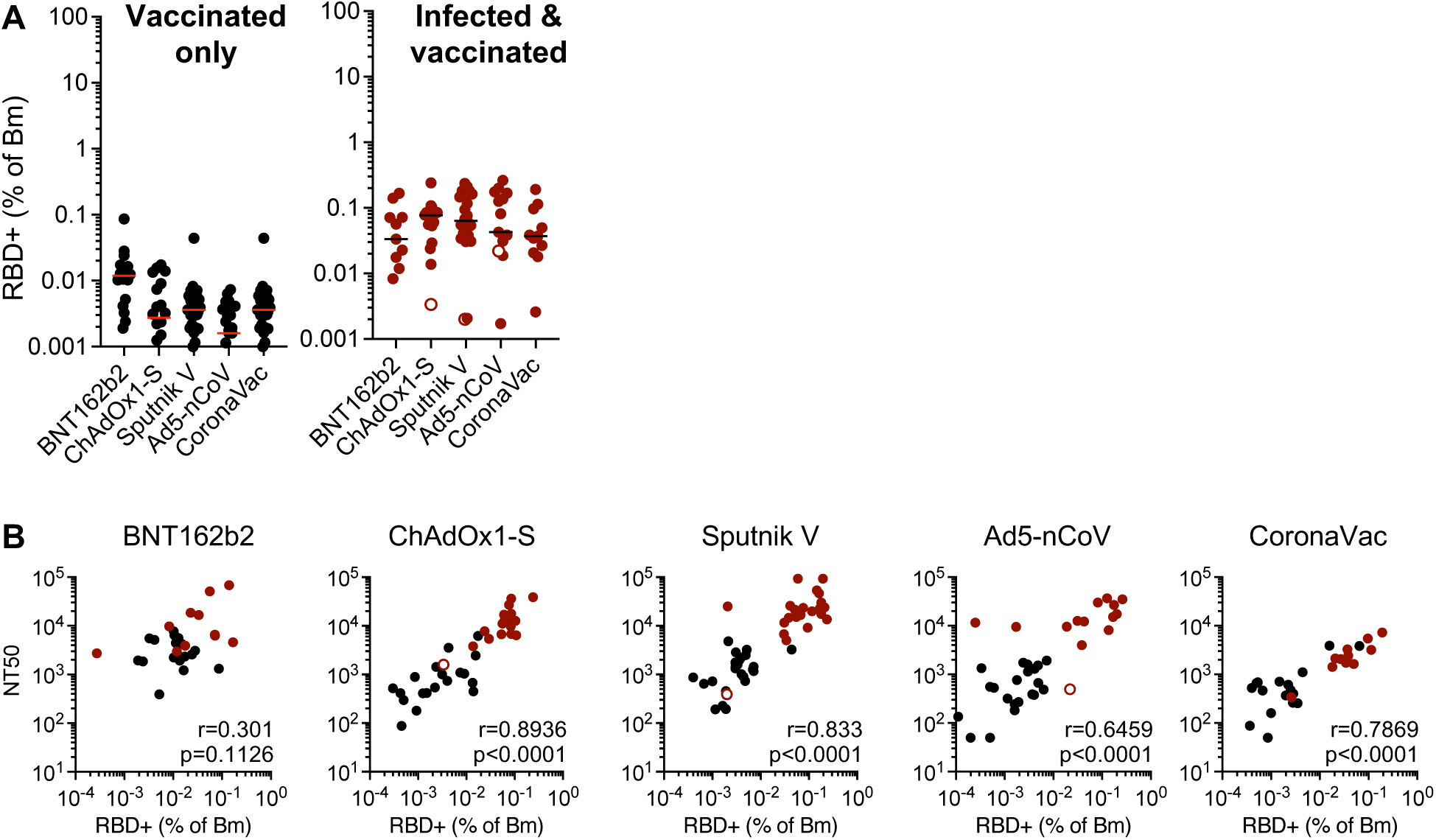
Quantification of RBD-specific B cells. RBD-specific memory B cells (Bm) in recipient PBMC were enumerated using FACS and a recombinant RBD fragment. **(A)** Percentage of S+ specific Bm cells that were specific for RBD in uninfected (left, black circles) or infected (right, red circles) vaccine recipients. Open red circles indicate samples from anti-N negative individuals as described in Fig. 2B. Horizontal lines indicated median percentage RBD+ (of Bm cells). **(B)** Correlation of neutralizing antibody titers (NT_50_) with percentage of RBD-binding Bm cells in each vaccine recipient group, colored as in (A). Spearmen’s correlation analysis r and p values are indicated.

**Supplementary Figure 5.**
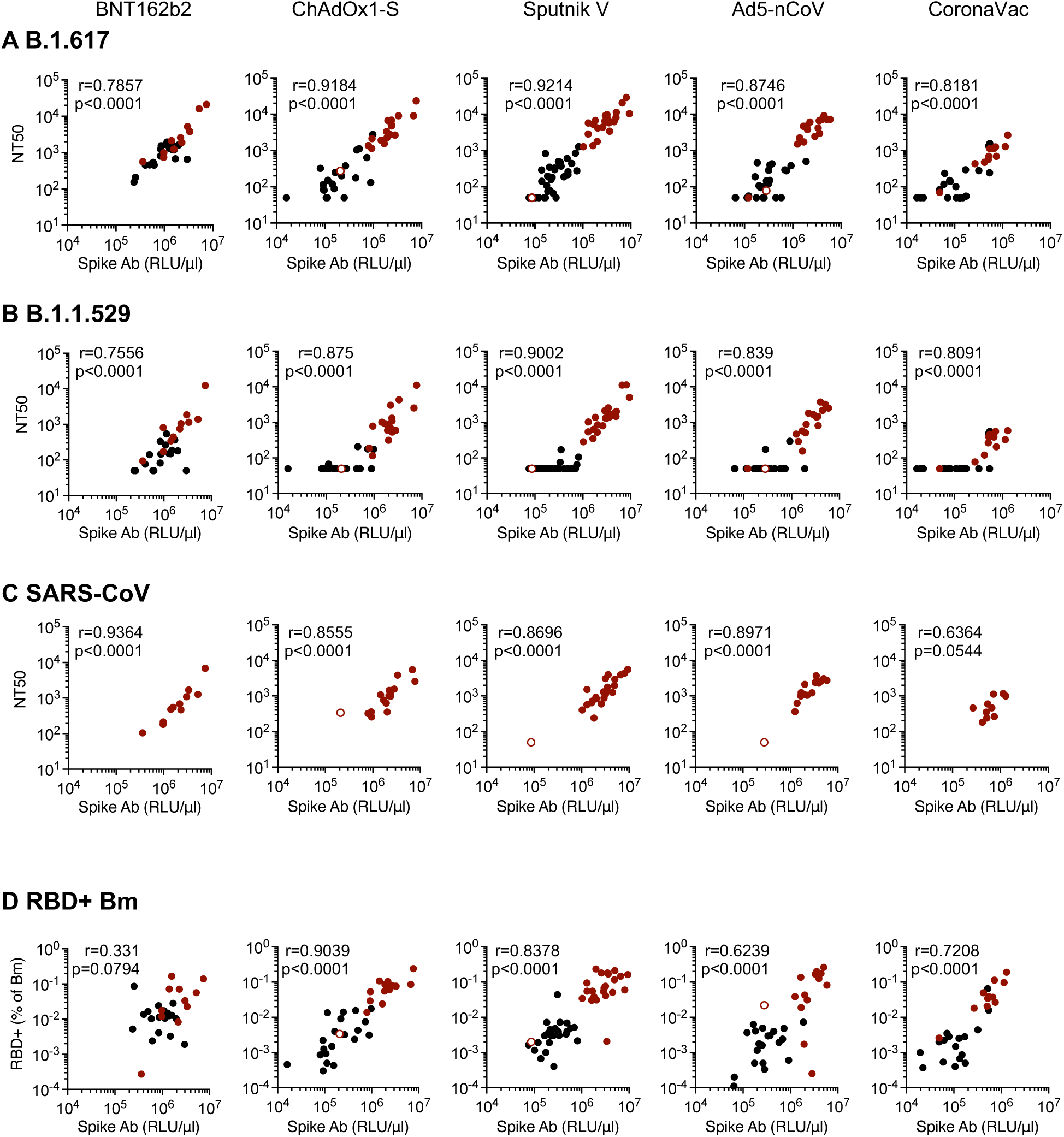
Correlation of Spike-binding antibodies with neutralization of SARS-CoV-2 variants, SARS-CoV and RBD-specific memory B cells. Correlation between Spike-binding antibodies (RLU/μl) with neutralizing antibody titers against **(A)** B.1.617.2(delta), **(B)** B.1.1.519(omicron) and **(C)** SARS-CoV. **(D)** Correlation between Spike-binding antibodies (RLU/μl) and percentage of memory B-cells that bind S RBD. Spearmen’s correlation analysis r and p values are indicated.

**Supplementary Figure 6.**
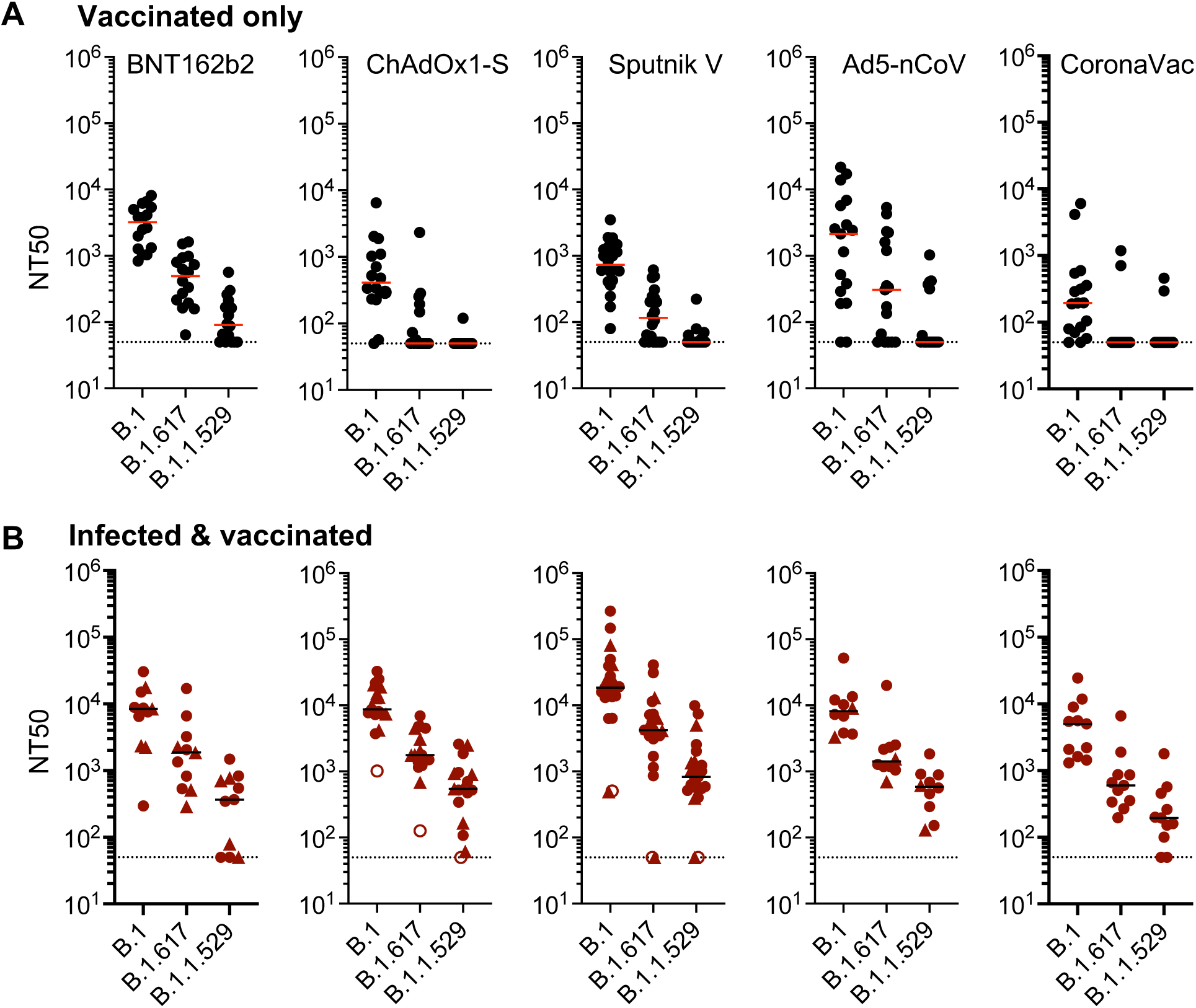
Plasma neutralization activity in second visit samples. NT_50_ values for plasmas from vaccine recipients collected 2.5-7.8 months after the first sample, against B.1 or other variants of concern as indicated. **(A)** Recipients had no documented prior infection with SARS-CoV-2. **(B)** Recipients were infected with SARS-CoV-2 either prior to the study as documented by a PCR positive test (closed circles) or at an unknown time prior or between vaccination and sample collection as confirmed by anti-N tests at the time of sample collection (closed triangles). Samples from participants with prior positive PCR tests but undetectable N titers in the first sample (open circles). The median of 2 independent experiments for each plasma is plotted. Dashed line indicates the lowest plasma dilution tested (1:50). Horizontal indicated median NT_50_ values.

